# Tiny Babies, Big Data: ICD Billing Code Patterns in Neonates Diagnosed with Genetic Disease in the Neonatal Intensive Care Unit

**DOI:** 10.64898/2026.02.08.26345857

**Authors:** Elly Brokamp, Retna Arun, Austin A. Antoniou, Bimal P. Chaudhari, Monica Wojcik

## Abstract

**Purpose:** Genetic diseases often present and are first diagnosed in the neonatal intensive care unit (NICU). Accurate identification of neonates with genetic diagnoses (GDs) in electronic health records (EHR) would enable a more complete understanding of their phenotypic spectrum, advancing care and personalized medicine. Prior research has used International Classification of Diseases (ICD) billing codes as proxies for GDs, though their accuracy for detecting confirmed GDs is uncertain. We evaluate the ICD codes for neonates with confirmed GDs and compare ICD billing code patterns between neonates with and without GD in two independent NICU cohorts.

**Methods:** Retrospective analysis of patients admitted to the Boston Children’s Hospital (BCH) level IV NICU (1,344 neonates) and Nationwide Children’s Hospital (NCH)’s neonatal network (33,315 neonates, mixed Level III/IV). For both cohorts, GDs captured by phecodes, aggregates of ICD codes, were compared with confirmed GDs. Two separate phenome-wide association studies (PheWAS) compared phecode patterns between neonates with GDs and those without, adjusting for sex, age at admission, gestational age, and NICU length of stay.

**Results:** Genetic phecodes were able to correctly identify 43.5% of neonates that received a GD in the BCH or NCH NICUs. Among 719 individuals with two or more genetic phecodes at BCH or NCH, 566 (78.72%) had a true GD. The BCH PheWAS analysis revealed a statistically significant positive association with atrioventricular septal defects and a negative association with bronchopulmonary dysplasia. The NCH pheWAS revealed 179 significantly associated phecodes, including many congenital anomalies.

**Conclusion:** The use of ICD codes to identify NICU infants with GDs is neither sensitive nor accurate, though phecode analysis demonstrated stronger accuracy than sensitivity. Our data highlight clinical features of NICU infants more commonly seen in those that receive a GD (congenital heart defects) and those that are not (BPD). Our results can help to better predict and identify NICU neonates that receive a GD.

## Introduction

There are ∼10,000 different rare diseases that affect ∼3-6% of individuals in the United States, with the vast majority being genetic in nature^1,2^. Most genetic diseases affect children and many conditions are associated with childhood mortality^3,4^. For those who live into adulthood, rare diseases necessitate high use of the healthcare system, accounting for almost half of the healthcare costs in the United States^5^. While only ∼5% of rare diseases currently have targeted approved treatments, in 2025 the Food and Drug Administration proposed a dedicated regulatory pathway for developing targeted rare genetic disease therapies^6,7^. These proposed pathways, along with research collaboratives, are quickly paving the path for an expansion of rare genetic disease therapeutics^8^. However, a major challenge facing rare disease clinical effectiveness research is the lack of large-scale data needed to garner impactful epidemiological insights-such as healthcare resource utilization, clinical signs and symptoms predictive of genetic disease, and morbidity and mortality rates and predictors. The ability to accurately identify individuals with genetic diseases in the electronic health records (EHR) is therefore crucial towards improving care and driving personalized medicine forward.

However, accurate and comprehensive identification of individuals in the EHR with genetic diagnoses (GDs) has continued to largely elude the bioinformatic field. Genetic testing results and corresponding GDs are not simple to identify at scale in the EHR, often requiring custom extraction and manual review at individual medical centers^9^. An natural language processing (NLP)-based approach, MonoMiner, has demonstrated high precision for the individuals with gene-based disease it identifies, but still has less than a 50% recall rate, omitting more than half of individuals with a GD^10^. Another EHR-phenotyping algorithm, PheIndex, is relatively sensitive (90% recall rate) at using criteria such as heart surgeries and specialist visits to identify those who have a genetic disorder or are at a high risk for a genetic disorder, but cannot differentiate between specific GDs^11^. In a simpler approach that also permits potential analysis of genetic diagnostic impact from administrative (billing) data alone, several research groups have used International Classification of Disease (ICD) billing codes as proxies for GD, although it is unclear how accurate ICD billing codes are in identifying those with confirmed molecular diagnoses-particularly in the neonatal intensive care unit (NICU), where phenotyping is more challenging and often incomplete^12^.

Due to their complex and often multisystem health needs, neonates admitted to the NICU often undergo diagnostic genetic testing compared to other clinical contexts^13^. Over the last decade, exome and genome sequencing (ES/ GS) have become more common as the first line of diagnostic genetic testing performed in the NICU, yielding a GD in 30-40% of critically ill neonates who undergo such testing^14,15,16,17^. However, optimal use of ES/GS in the NICU remains incompletely defined, and accurate evaluation of clinical outcomes following testing and subsequent GD would help healthcare leaders to make informed decisions of when to offer such testing^18,19,20^. We therefore evaluated the capabilities of ICD 9/10 billing codes to identify infants with a GD and then we compare ICD code patterns between neonates with and without GD in two independent NICU cohorts.

## Methods

### NICU cohorts

Boston Children’s Hospital (BCH)’s NICU dataset includes 1,389 neonates who have been admitted to BCH’s Level IV NICU in Boston, Massachusetts from 2011 to 2025 who underwent genetic evaluations. The BCH NICU database is organized through a Research Electronic Data Capture (REDCap) databse system and is manually curated from the EHR by a team of trained research assistants^21^. Abstracted data include demographics, clinical information such as presenting phenotype, and genetic testing information such as type of test sent and diagnostic outcome. Infant date of birth, sex, birthweight, gestational age at birth, age at NICU admission, length of NICU admission, date of genetic consult, genetic tests ordered, available genetic testing results, genetic diagnoses, and clinical outcomes at one year of life are recorded. The BCH NICU REDCap password protected that only affiliated BCH clinicians and researchers have access to it.

Data from Nationwide Children’s Hospital (NCH)’s Neonatal Network database includes 33,315 neonates who were admitted to the NICU from 2010-2021. The NCH Neonatal Network consists of six Level III NICUs and one level IV NICU all in central Ohio, but only data from the Level IV (9,443 neonates) were used for our analyses. Clinical and administrative data on admitted infants are collected into a single Research Data Warehouse (RDW). Each infant’s birthweight, gestational age at birth, available laboratory testing results, consults from genetic providers, ICD9/10 billing codes, the age an ICD9/10 billing code was used, and the length of NICU admission are recorded in the RDW. The captured laboratory tests are defined as either genetic (including ES/GS, next-generation sequencing (NGS), microarrays, karyotypes, Flourescent In Situ Hybridization, methylation, repeat expansion, and uniparental disomy testing) or non-genetic. Patients are given a unique identifier, allowing linkage of data from delivery centers with data from NCH Level IV NICU, facilitating review of antenatal screening and testing outcomes.

Neonates that received a documented GD during their NICU admission are cases (as opposed to using genetic phecodes to indicate a diagnosis) and neonates who did not receive a genetic diagnosis or a genetic phecode are controls. BCH’s documented GDs are manually curated in their NICU database and those without a documented GD are considered controls. Patients in the NCH cohort were labelled as having a GD if they either had a genetic test with a molecular diagnosis in the chart or confirmed genetic diagnosis described by a medical geneticist (even in the study team did not have access ot the actual test result). Those that did not receive positive genetic testing results or a specifically indicated negative ES/GS are controls for the NCH cohort.

### Statistical analyses

We use phecodeX phecodes to systematically group ICD9/10 billing codes during NICU admissions at both children’s hospitals into meaningful aggregates^22^. For BCH neonates, GDs indicated by phecodes in the genetic disease (GE_) chapter were compared with manually confirmed GDs in the BCH NICU database. For these analyses, we required two instances of a GE_ phecode to indicate a GD captured by phecodes. A member of our team manually investigated both the accuracy of genetic phecodes and the recall rate of genetic phecodes by comparing the manually curated GDs in the RedCAP database with GD phecodes. We also indicated whether a genetic phecode, while technically correct, has a more specific, but still accurate genetic phecode available. For example, a neonate with a confirmed GD of Trisomy 21 with phecode GE_960 “Chromosomal anomalies” is technically correct, but the phecode GE_960.11 “Down syndrome” would be more specific. We reviewed the cohort for GDs up to one year of life. For the NCH cohort, which does not have the specific GD documented, we compared how many neonates with a GD documented in their database were also indicated to have a GD by phecodes.

We also used phenotype-wide association studies (PheWAS) to compare phecode patterns between neonates that received a confirmed GD during their NICU admission (cases) and those that did not (controls) using the PheWAS R package^23^. PhecodeX mapping was used to aggregate ICD9/10 billing codes into phecodes for the billing codes received during a neonate’s NICU admission^22^.

PheWAS analyses are performed using multiple logistical regressions for all available phecodes with the regression models adjusting for sex, age at NICU admission, gestational age, and the length of stay in the NICU. We limited analyses to non-genetic phecodes with at least 15 individuals, resulting in 591 regressions for different phecodes in the BCH analysis and 849 regressions for different phecodes in the NCH analysis. Based on the number of regressions for each tested phecode, the expected Bonferonni p-value thresholds are 8.6x10^-5^ (BCH) and 5.9x10^-5^ (NCH).

## Results

### NICU populations

There are 197 neonates who received a GD in the BCH NICU and 1,118 who did not receive a GD during their NICU stay. There are 885 neonates who received a GD in the NCH NICU database and 8,558 who did not receive a GD during their NICU stay. Demographics and clinical data, including genetic diagnosed status, of both cohorts are listed in **Table 1**. Consistent with previously observed trends, there are slightly more males than females in both NICU cohorts^24^. There is not a significant difference between gestational age between those that received a GD and those that did not in either BCH or NCH NICU cohort.

**Table 1.**
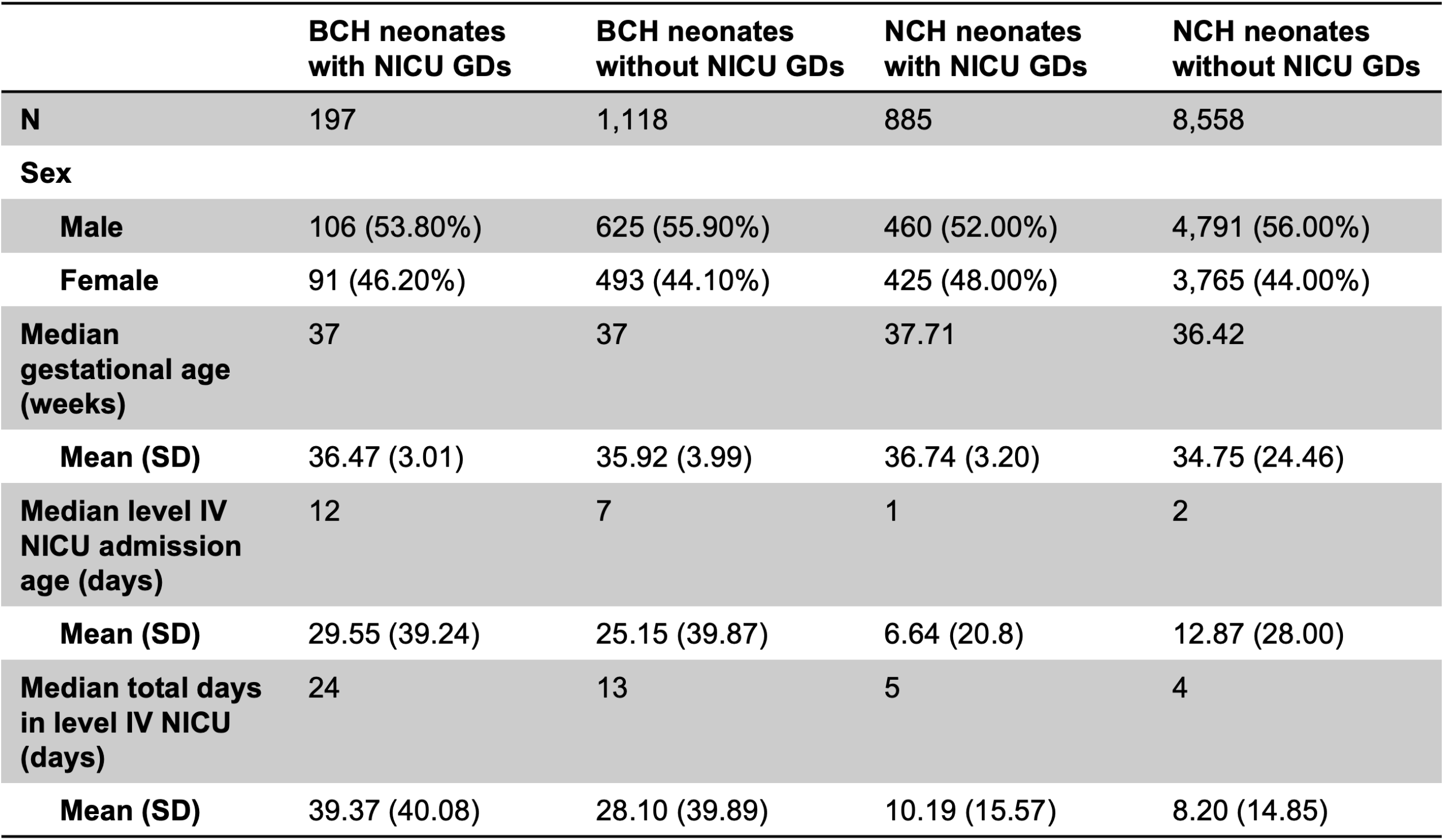
NICU cohort cases and controls demographics.

### Genetic phecodes’ capture of confirmed genetic diagnoses

The BCH NICU cohort includes 388 neonates who received a GD within one year of life. Of those diagnosed within one year of life, 42 were diagnosed before their first NICU admission, 197 were diagnosed during their NICU admission, and 149 were diagnosed after discharge from the NICU. Of the 197 neonates who received a GD during their NICU admission, genetic phecodes generate using ICD9/10 codes from this NICU admission were able to correctly identify 59 (30.0%).

Considering diagnoses beyond the NICU timepoint, among 152 neonates captured by genetic phecodes in the BCH NICU cohort, 126 (82.9%) correctly corresponded to their documented genetic diagnosis. ICD billing codes, and thus phecodes, are organized in a tiered structure. So, while some individuals receive a GD phecode that is technically correct, there is another phecode that is more specific, and more appropriate, to the documented genetic diagnosis. Of those with a correct genetic phecode, there are 16 (12.7%) that are technically correct, but that could be more specific. This concept is highlighted by five individuals who received the generic parent phecode “GE_960 Chromosomal anomalies”, but there are more specific genetic phecodes that correspond to their diagnoses. For example, an individual with a 16q deletion would be more appropriately described by “GE_960.28 Other chromosome microdeletions” rather than the generic “GE_960 Chromosomal anomalies”.

Genetic phecodes incorrectly classified 25 (16.45%) neonates as having a GD, with almost half (11) not having a documented genetic diagnosis at all. Further chart review found that 7 of the individuals without a documented genetic diagnosis had abnormal newborn screening (such as concern for Very Long Chain Acyl-CoA Dehydrogenase Deficiency on newborn screening) but received a phecode indicating an ultimately incorrect diagnosis (“GE_962.31 Long chain/very long chain acyl CoA dehydrogenase deficiency”). The remaining individuals have had genetic diagnoses, but the incorrect genetic phecode was given. For example, an infant may have a phecode indicating Trisomy 21 (“GE_960.11 Down syndrome”) but their actual genetic diagnosis in the BCH NICU database is Noonan syndrome. **Figure 1** demonstrates a Sankey plot of how many individuals were correctly captured by genetic phecodes.

**Figure 1.**
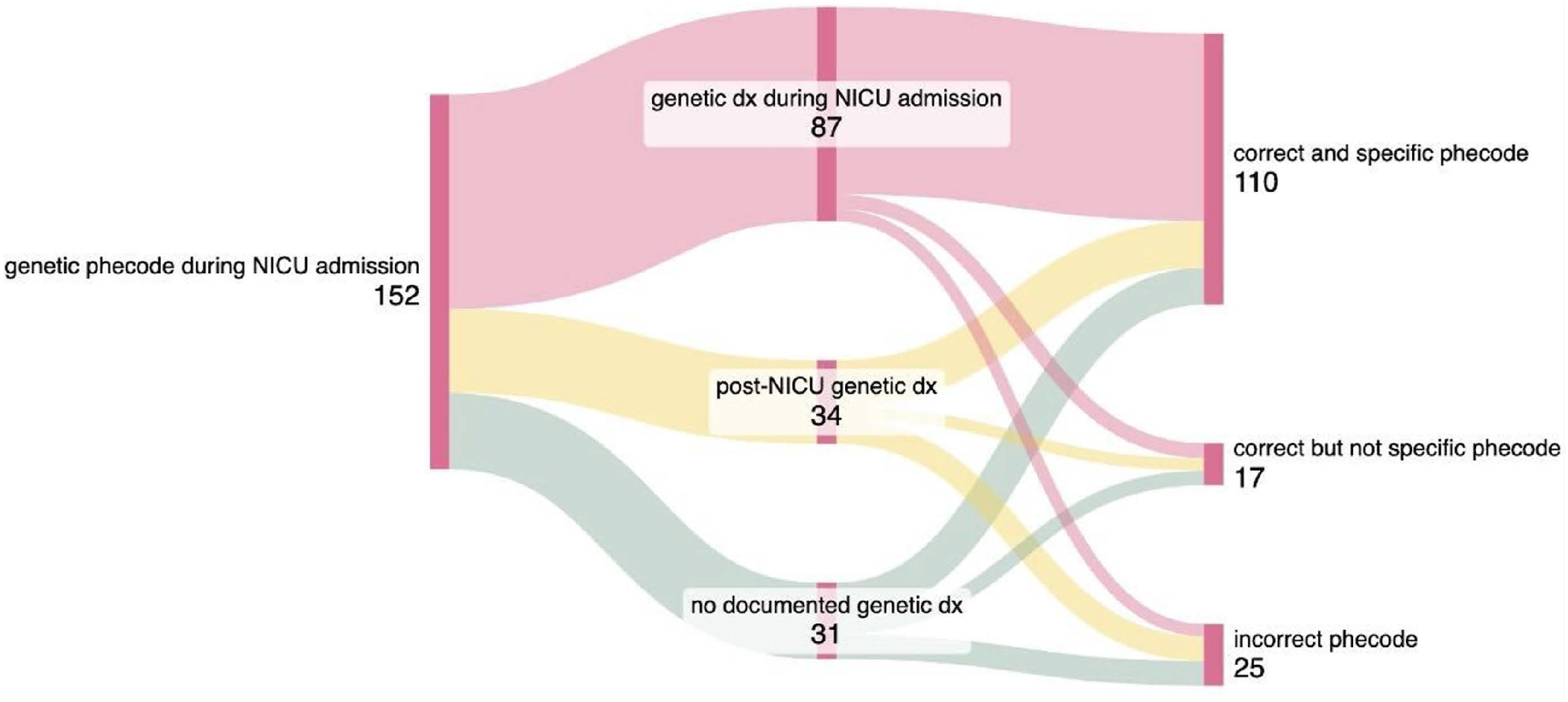
NICU cohort cases and controls demographics. A Sankey plot demonstrating how many individuals who had a genetic diagnosis indicated by phecodes had a the correct and most specific genetic phecode, a correct but not specific genetic phecode, or an incorrect genetic phecode when compared to their manually captured true genetic diagnoses.

In the NCH cohort, out of the 885 neonates with a confirmed GD, 439 (49.60%) received a billing code indicative of a GD. Out of the 567 individuals who received genetic phecodes, 128 (22.57%) did not have a GD in the database (indicated by a positive genetic test). **Table 2** summarizes genetic phecodes capabilities of identifying those with a true GD in both cohorts.

**Table 2.**
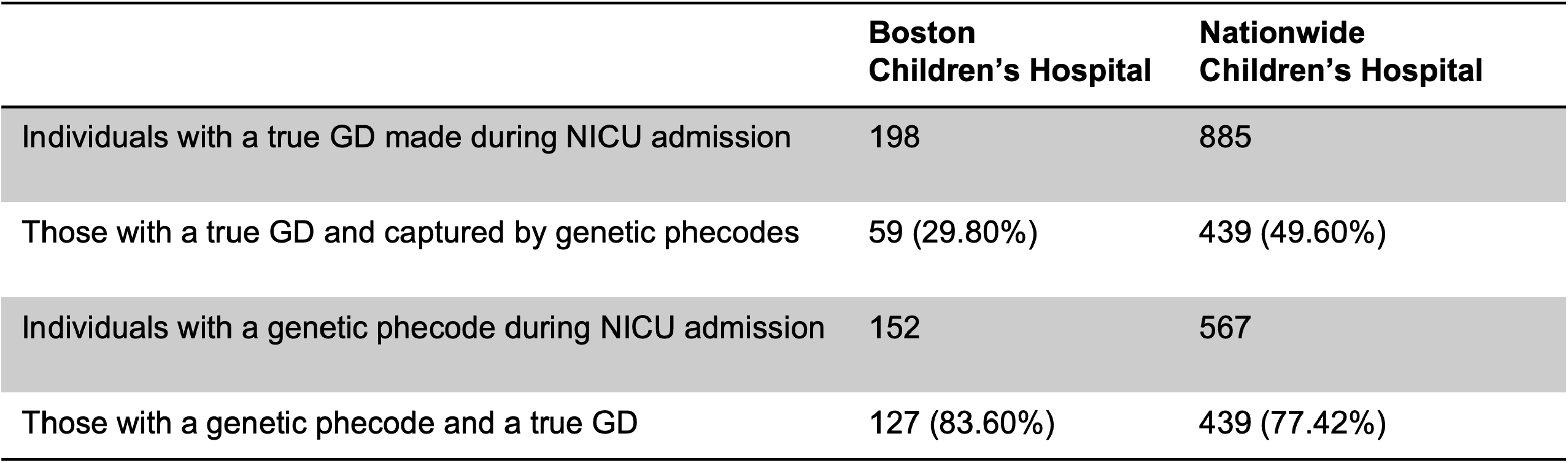
Recall rate and positive predictive value of genetic phecodes.

### NICU genetic diagnosis pheWAS

In addition to clarifying the accuracy of ICD10 codes to classify infants with a GD in the NICU, we also sought to better understand the possible ICD9/10 billing code patterns for individuals who received a GD during their NICU admission, compared to those who did not receive a GD, as this could help predict the likelihood of GD in the NICU based on phenotypic features. For this pheWAS analysis, we used both the BCH and NCH NICU populations. Both pheWAS analyses demonstrated phecodes significantly associated with neonates who received a GD during their NICU admission.

The BCH pheWAS revealed two phecodes that passed the Bonferroni-corrected threshold (p=8.6x10^-5^). Atrioventricular septal defects (AVSDs) are significantly (p =2.93x10^-5^) positively associated (beta=1.52) with neonates who received a GD in the NICU. Bronchopulmonary dysplasia (BPD) is significantly (p =6.56x10^-5^) negatively associated (beta= -1.87) with individuals who received a GD in the NICU. **Figure 2**, a Manhattan plot, demonstrates phecodes that were either associated or dis-associated with receiving a GD in the BCH NICU. **Supplementary Table 1** contains complete BCH pheWAS results.

**Figure 2.**
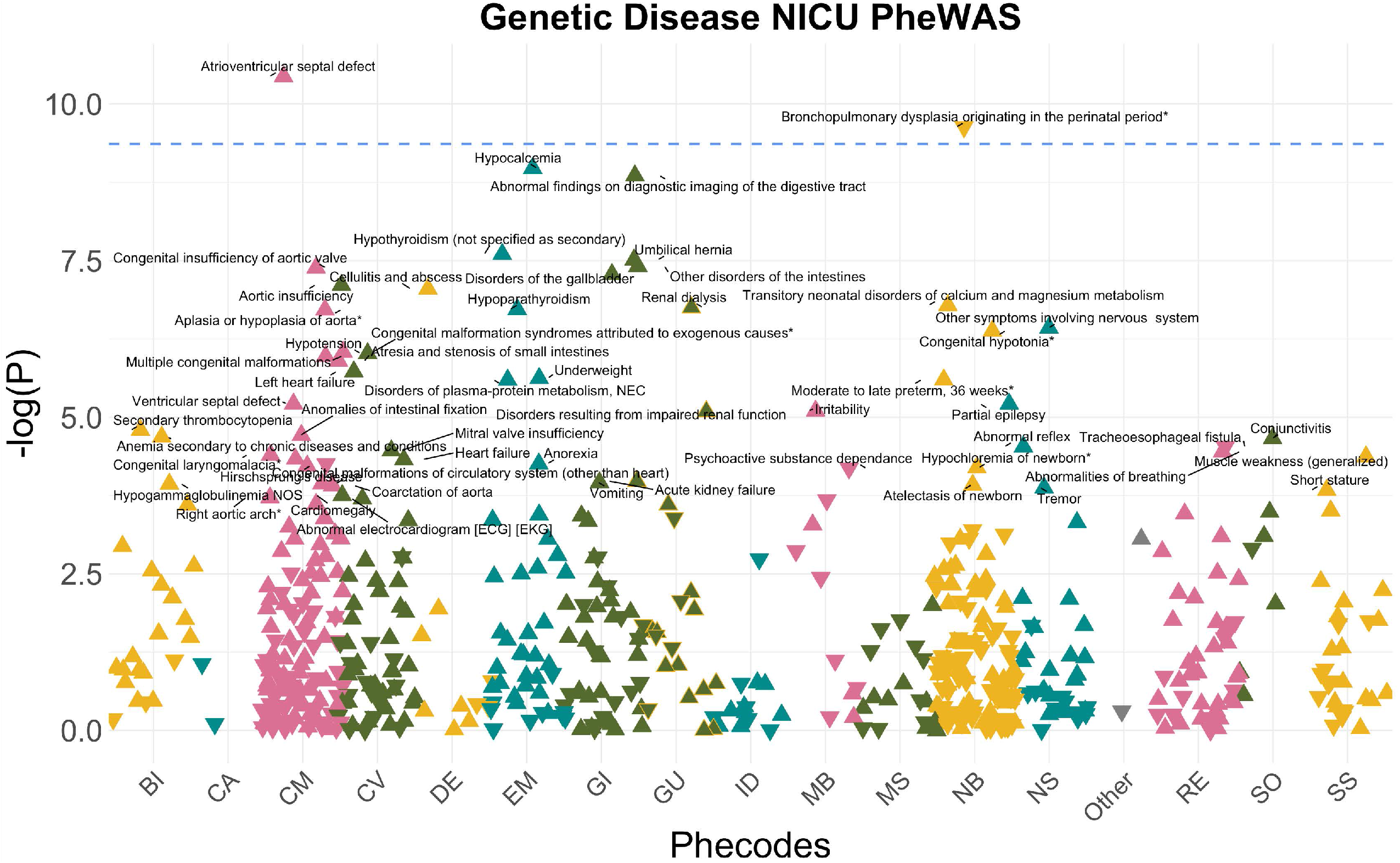
PheWAS plot of neonates with genetic diagnosis during BCH NICU admission. A pheWAS plot of those with confirmed genetic diagnoses in the BCH NICU. PhecodeX chapters are categorized by color along the x-axis (Bl= Blood Immune, CA= Cancer, CV= Cardiovascular, CM= Congenital Malformations, DE= Dermatological, EM= Endocrine Metabolic, GI= Gastrointestinal, GE= Genetic, GU= Genitourinary, ID= Infections, MB= Mental, MS= Musculoskeletal, NB= Neonatal, NS= Neurological, RE= Respiratory, SO= Sensory Organs, SS= Symptoms). The blue dotted line indicates the significance threshold of p <= p=8.6x10^-5^. Triangle plot points pointing up indicate a positive association and those pointing down indicate a negative association with GD.

The NCH pheWAS revealed 179 phecodes significantly associated (p< 2.6x10^-5^) with receiving a GD in the NCH NICU. AVSDs are also significantly associated with NICU GD in NCH’s cohort (p=3.06x10^-23^, beta=2.49), along with similar congenital heart defects (CHDs): ventricular septal defects (VSDs; p=1.53x10^-67^, beta=2.09), patent ductus arteriosus (PDAs; p= 7.44x10^-62^, beta=1.35), and atrial septal defects (ASDs; p= 1.1x10^-48^, beta=1.10). Phecodes within the congenital malformation chapter beyond CHDs revealed the top positive associations with GDs, including: multiple congenital anomalies (MCA; p = 1.42x10^-65^, beta=3.36), other congenital malformations (p=1.05x10^-41^, beta=2.15), congenital deformities of skull, face, and jaw (p=1.89x10^-36^, beta=2.06), cleft palate (p=1.31x10^-28^, beta=2.48), congenital deformities of feet (p=5.65x10^-24^, beta=2.00), and congenital anomalies of limbs (p=4.98x10^-21^, beta=2.49). Two phecodes were significantly dis-associated with GD diagnosis: noxious substances (p=3.0x10^-12^, beta= -0.91) and hypoxic ischemic encephalopathy (p=4.4x10^-5^, beta= -0.79). **Figure 3** displays the NCH NICU GD pheWAS Manhattan. **Supplementary Table 2** contains complete NCH pheWAS results. **Table 3** demonstrates the strongest positively and negatively associated phecodes with GD in both the BCH and NCH NICU cohorts.

**Table 3.**
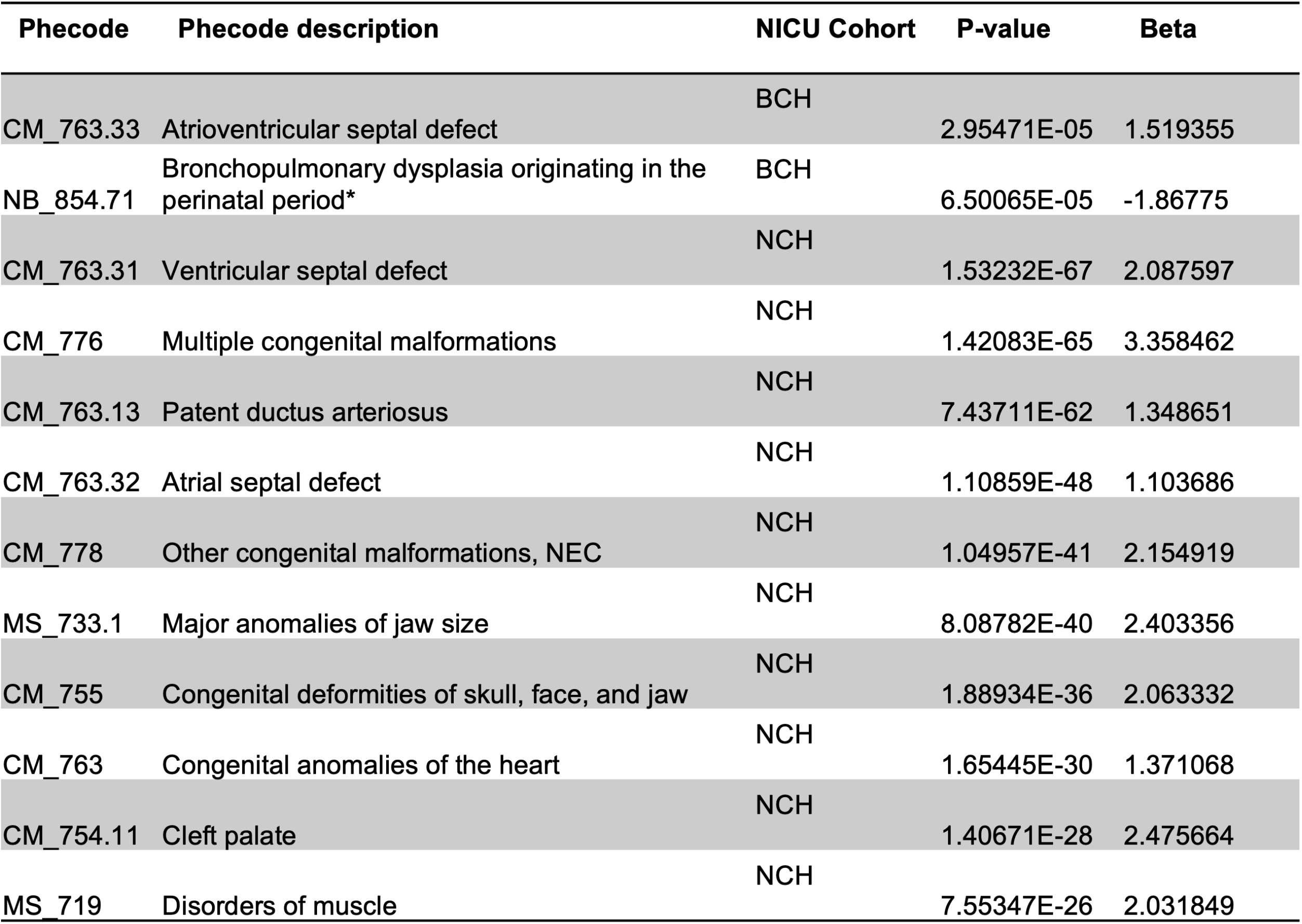
Top positively and negatively associated phecodes in those with NICU genetic diagnosis.

**Figure 3.**
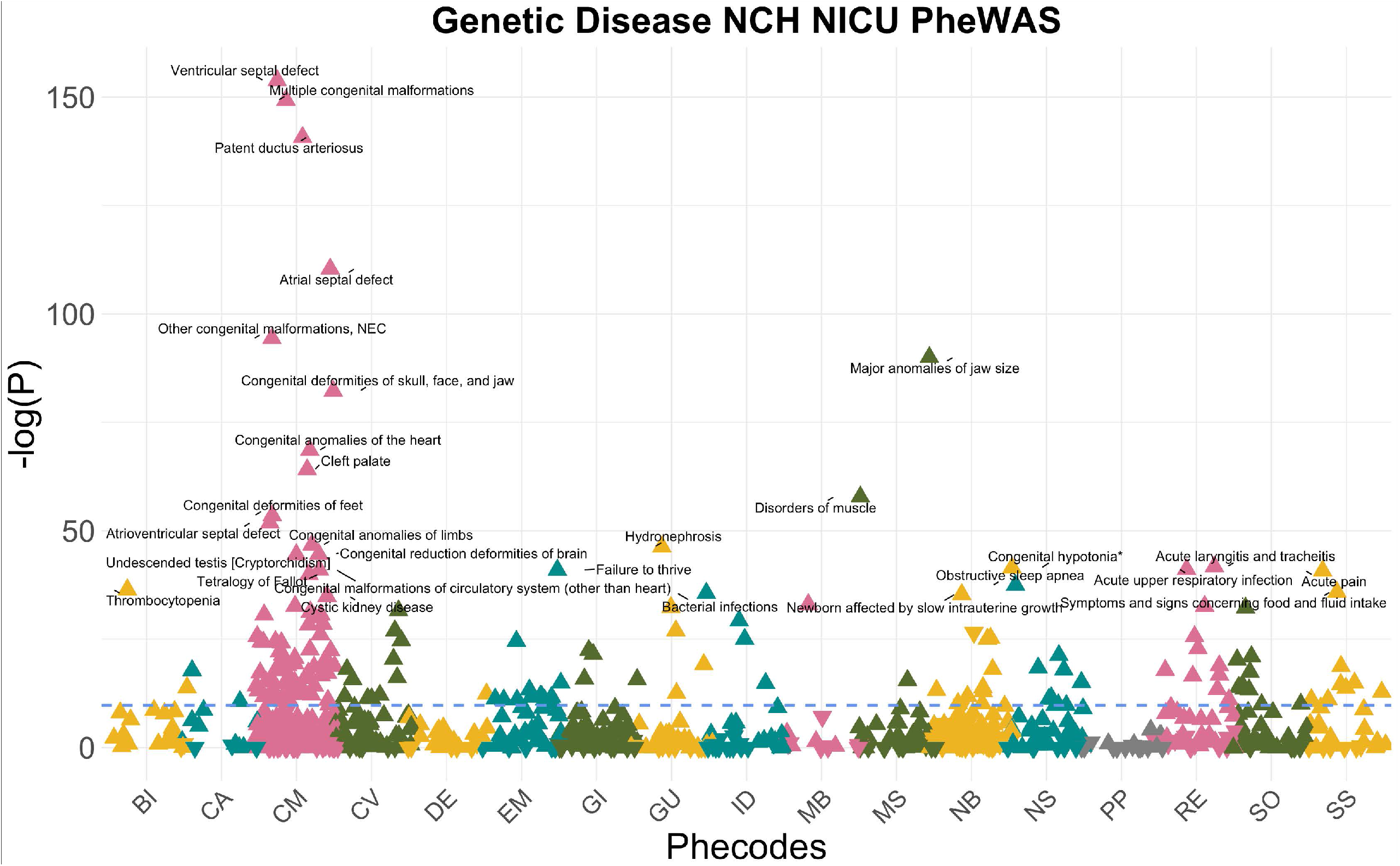
pheWAS of neonates with genetic diagnosis during NCH NICU admission. A pheWAS of those with confirmed genetic diagnoses in the NCH NICU. PhecodeX chapters are categorized by color along the x-axis. The blue dotted line indicates the significance threshold of p <= p=2.6x1^5^. Triangle plot points pointing up indicate a positive association and those pointing down indicate a negative association with GD.

## Discussion

Due to their care of complicated and acute presentations, NICU populations have been found to generally have a higher rate of individuals with genetic disease (10-50% depending on how many neonates are evaluated and the genetic technology used) as compared to the general population^25,26^. The NICU admission also serves as one of the earliest time points researchers can begin to categorize and better understand the phenotypes of rare genetic diseases. As neonatal genomic medicine accelerates, rare disease clinical effectiveness and outcomes research would greatly benefit from developing an accurate and comprehensive method to retrospectively identify neonates affected by GDs that does not require manually curated databases. In addition, this same methodology could potentially be applied prospectively at the point of NICU admission to select infants who stand to benefit the most from diagnostic genetic testing (ES/GS).

Towards the first goal, our phecode analysis of BCH NICU data revealed that the use of ICD9/10 codes to identify NICU infants with GDs has neither a high recall rate nor accuracy, though phecode analysis demonstrated stronger accuracy than recall rate. The phecode method’s recall rate (∼3.5%) for identifying neonates with a GD made in the NICU was lower than previously reported in a similar analysis using hospital-side data (53%)^10^. Conversely, genetic phecodes in the NICU population had a similar accuracy rate (83.6%) than an NLP method identifying those with confirmed GDs throughout an entire hospital system (87%)^10^. This highlights challenges in phenotyping NICU infants, who often present with non-specific symptoms or incomplete manifestations of the underlying genetic conditions. In addition, we acknowledge that the ICD billing code system is designed to classify common diseases, leaving most genetic diseases without a matching ICD billing code. But even when there is a more specific ICD 9/10 billing code appropriate to a patient’s diagnosis, we found that the more specific code was not used 12.7% of the time in our NICU cohort. Less often, neonates received a genetic phecode for the wrong GD (11.1%) and neonates without a GD at all occasionally would be classified as having a GD via a genetic phecode (5.3%). Thus, relying on administrative data – ICD billing codes – to classify infants with GD for the purposes of clinical epidemiological investigations has crucial shortfalls which may lead to biased conclusions.

Towards the second goal of prospective identification of infants with GD to direct testing practices, our pheWAS data highlight features of NICU infants more commonly seen in those that receive a GD than infants who did not receive a GD. In the smallest cohort, BCH infants who received a GD in the NICU were significantly more likely to have an AVSD. This significant association may be due to the common chromosomal anomaly diagnosis, Trisomy 21, but there are also other minor CHDs (e.g ventricular septal defects, patent ductus arteriosus (PDA), atrial septal defects) that have a significant association with neonates that received GDs in NCH’s cohort. Interestingly, PDAs are most common in premature neonates, but there are several Mendelian syndromes associated with PDAs and analyses were corrected for gestational age^27^.The only significantly dis-associated phecode in the BCH’s cohort, BPD is rarely associated with genetic diseases^28^. However, BPD was mildly associated (p=0.0022, beta=0.59) with GDs in the NCH cohort. This is likely an artifact of the fact that severe BPD due to complications of prematurity but without complex, multi-system involvement complicating BPD Comprehensive Care Center, which medically complex neonates, including those more likely to have GDs, who also have BPD remain in the main Level IV NICU, thus biasing the NCH dataset with regard to how babies with BPD are represented in the data. Not surprisingly, congenital anomalies were highly associated with GDs in both cohorts.

We recognize that ICD billing codes are not a perfect indication of disease. This sentiment is especially true for genetic disease ICD billing codes. Other phenotypes, such as congenital anomalies, can be identified by ICD billing codes with better accuracy^29^. Besides the peaks of significantly associated congenital anomalies, other results also make clinical sense, such as GD’s positive association with congenital hypotonia (NCH p= 9.62X10^-19^, beta = 1.70 ; BCH p= 2.71 X10^-3^, beta = 0.644), which is included in the phenotypic description of fifty genetic diseases in OMIM^30^. The limitations in our pheWAS analyses are indicative of limitations of EHR-based research in general, such as inappropriate billing codes being given or incomplete capture of phenotypes; we argue that our goal of understanding billing code patterns of those with GDs circumvents these limitations given our interest in “real-life” billing code data.

Despite these limitations, our pheWAS results for individuals that receive a GD while in the NICU has the potential to help better predict future NICU neonates that will receive a GD. Many efforts have been made to use EHR data to select individuals that would benefit from a genetics consult or genetic testing^11,31^, but to our knowledge none have been restricted to the NICU setting. This early timepoint of care for some of the sickest patients is one that would benefit from targeted genetics care. As genetic disease therapeutics become more available, identifying those who are eligible for treatment will be invaluable. Future expanded analyses to additional hospitals can help build a NICU-specific GD prediction model.

## Data availability

All data produced in the present work are contained in the manuscript

## References

1. Nguengang Wakap S, Lambert DM, Olry A, et al. Estimating cumulative point prevalence of rare diseases: analysis of the Orphanet database. Eur J Hum Genet. 2020;28(2):165–173. doi:10.1038/s41431-019-0508-0

2. Haendel M, Vasilevsky N, Unni D, et al. How many rare diseases are there? Nat Rev Drug Discov. 2020;19(2):77–78. doi:10.1038/d41573-019-00180-y

3. Wojcik MH, Schwartz TS, Thiele KE, et al. Infant mortality: the contribution of genetic disorders. J Perinatol Off J Calif Perinat Assoc. 2019;39(12):1611–1619. doi:10.1038/s41372-019-0451-5

4. Shen T, Lee A, Shen C, Lin CJ. The long tail and rare disease research: the impact of next-generation sequencing for rare Mendelian disorders. Genet Res. 2015;97:e15. doi:10.1017/S0016672315000166

5. Navarrete-Opazo AA, Singh M, Tisdale A, Cutillo CM, Garrison SR. Can you hear us now? The impact of health-care utilization by rare disease patients in the United States. Genet Med. 2021;23(11):2194–2201. doi:10.1038/s41436-021-01241-7

6. Committee on Diagnostic Error in Health Care, Board on Health Care Services, Institute of Medicine, The National Academies of Sciences, Engineering, and Medicine. Improving Diagnosis in Health Care. (Balogh EP, Miller BT, Ball JR, eds.). National Academies Press; 2015:21794. doi:10.17226/21794

7. FDA’s New Plausible Mechanism Pathway | New England Journal of Medicine. Accessed January 30, 2026. https://www.nejm.org/doi/full/10.1056/NEJMsb2512695

8. Belgrad J, McConnell E, Leonard S, et al. The N=1 Collaborative: advancing customized nucleic acid therapies through collaboration and data sharing. Nucleic Acids Res. 2025;53(8):gkaf346. doi:10.1093/nar/gkaf346

9. Bastarache L, Tinker RJ, Schuler BA, et al. Characterizing trends in clinical genetic testing: A single-center analysis of EHR data from 1.8 million patients over two decades. Am J Hum Genet. 2025;112(5):1029–1038. doi:10.1016/j.ajhg.2025.03.009

10. Wu DW, Bernstein JA, Bejerano G. Discovering monogenic patients with a confirmed molecular diagnosis in millions of clinical notes with MonoMiner. Genet Med. 2022;24(10):2091–2102. doi:10.1016/j.gim.2022.07.008

11. Webb BD, Lau LY, Tsevdos D, et al. An algorithm to identify patients aged 0–3 with rare genetic disorders. Orphanet J Rare Dis. 2024;19:183. doi:10.1186/s13023-024-03188-9

12. Gonzaludo N, Belmont JW, Gainullin VG, Taft RJ. Estimating the burden and economic impact of pediatric genetic disease. Genet Med Off J Am Coll Med Genet. 2019;21(8):1781–1789. doi:10.1038/s41436-018-0398-5

13. Callahan KP, Radack J, Wojcik MH, et al. Hospital-level variation in genetic testing in children’s hospitals’ neonatal intensive care units from 2016 to 2021. Genet Med. 2023;25(3):100357. doi:10.1016/j.gim.2022.12.004

14. D’Gama AM, Rosario MCD, Bresnahan MA, Yu TW, Wojcik MH, Agrawal PB. Integrating rapid exome sequencing into NICU clinical care after a pilot research study. NPJ Genomic Med. 2022;7:51. doi:10.1038/s41525-022-00326-9

15. Genome sequencing as a first-line diagnostic test for hospitalized infants. Genet Med. 2022;24(4):851–861. doi:10.1016/j.gim.2021.11.020

16. Callahan KP, Mueller R, Flibotte J, Largent EA, Feudtner C. Measures of Utility Among Studies of Genomic Medicine for Critically Ill Infants. Accessed November 25, 2025. https://jamanetwork.com/journals/jamanetworkopen/fullarticle/2794989

17. Pandey R, Brennan NF, Trachana K, et al. A meta-analysis of diagnostic yield and clinical utility of genome and exome sequencing in pediatric rare and undiagnosed genetic diseases. Genet Med. 2025;27(6):101398. doi:10.1016/j.gim.2025.101398

18. Ziegler A, Koval-Burt C, Kay DM, et al. Expanded Newborn Screening Using Genome Sequencing for Early Actionable Conditions. JAMA. 2025;333(3):232–240. doi:10.1001/jama.2024.19662

19. Reimers R, Bailey M, Brown C, et al. Clinical utility and cost-effectiveness of BeginNGS newborn screening by genome sequencing and standard newborn screening for severe childhood genetic diseases: an adaptive, international and comparative clinical trial. BMJ Open. 2025;15(11):e098609. doi:10.1136/bmjopen-2024-098609

20. Lunke S, Downie L, Caruana J, et al. Feasibility, acceptability and clinical outcomes of the BabyScreen+ genomic newborn screening study. Nat Med. 2025;31(12):4236–4245. doi:10.1038/s41591-025-03986-z

21. Frelich MJ, Bosler ME, Gould JC. Research Electronic Data Capture (REDCap) electronic Informed Consent Form (eICF) is compliant and feasible in a clinical research setting. Int J Clin Trials. 2015;2(3):51–55. doi:10.18203/2349-3259.ijct20150591

22. Shuey MM, Stead WW, Aka I, et al. Next-generation phenotyping: introducing phecodeX for enhanced discovery research in medical phenomics. Bioinforma Oxf Engl. 2023;39(11):btad655. doi:10.1093/bioinformatics/btad655

23. Carroll RJ, Bastarache L, Denny JC. R PheWAS: data analysis and plotting tools for phenome-wide association studies in the R environment. Bioinforma Oxf Engl. 2014;30(16):2375–2376. doi:10.1093/bioinformatics/btu197

24. Kim Y, Ganduglia-Cazaban C, Chan W, Lee M, Goodman DC. Trends in neonatal intensive care unit admissions by race/ethnicity in the United States, 2008–2018. Sci Rep. 2021;11:23795. doi:10.1038/s41598-021-03183-1

25. Swaggart KA, Swarr DT, Tolusso LK, He H, Dawson DB, Suhrie KR. Making a Genetic Diagnosis in a Level IV Neonatal Intensive Care Unit Population: Who, When, How, and at What Cost? J Pediatr. 2019;213:211–217.e4. doi:10.1016/j.jpeds.2019.05.054

26. Kingsmore SF, Cole FS. The Role of Genome Sequencing in the NICU. Annu Rev Genomics Hum Genet. 2022;23:427–448. doi:10.1146/annurev-genom-120921-103442

27. Lewis TR, Shelton EL, Van Driest SL, Kannankeril PJ, Reese J. Genetics of the patent ductus arteriosus (PDA) and pharmacogenetics of PDA treatment. Semin Fetal Neonatal Med. 2018;23(4):232–238. doi:10.1016/j.siny.2018.02.006

28. Torgerson DG, Ballard PL, Keller RL, et al. Ancestry and genetic associations with bronchopulmonary dysplasia in preterm infants. Am J Physiol - Lung Cell Mol Physiol. 2018;315(5):L858–L869. doi:10.1152/ajplung.00073.2018

29. Brokamp E, Miller-Fleming T, Scalici A, et al. Systematic method for classifying multiple congenital anomaly cases in electronic health records. Genet Med Off J Am Coll Med Genet. 2025;27(6):101415. doi:10.1016/j.gim.2025.101415

30. Entry Search - “congenital hypotonia” - OMIM - (OMIM.ORG). Accessed February 3, 2026. https://www.omim.org/search?index=entry&limit=10&sort=score+desc%2C+prefix_sort+desc&search=%22congenital+hypotonia%22

31. Morley TJ, Han L, Castro VM, et al. Phenotypic signatures in clinical data enable systematic identification of patients for genetic testing. Nat Med. 2021;27(6):1097–1104. doi:10.1038/s41591-021-01356-z

